# Bimodal distributions of anti-*Trypanosoma cruzi* antibody levels in blood donors are associated with parasite detection and antibody waning in peripheral blood

**DOI:** 10.1101/2024.11.26.24317961

**Authors:** Mirta C. Remesar, Ester C. Sabino, Lewis F. Buss, Claudio D. Merlo, Mónica G. López, Sebastián L. Humeres, Pavón A. Héctor, C. Clara Di Germanio, Sonia Bakkour Coco, Léa C. Oliveira-da Silva, Marcelo Martins Pinto Filho, Antonio Luiz P. Ribeiro, Michael P. Busch, Ana E. del Pozo

## Abstract

**Background:** In our previous study of blood donors in the Argentinian Chaco Province, we documented bimodal distributions of anti-*Trypanosoma cruzi* antibody (Ab) levels, suggesting potential self-cure in donors with low-reactive samples. This study aimed to correlate “high” and “low” Ab level groups, defined by a mathematical model, with parasitemia and electrocardiogram findings. Ab decline over time was also assessed.

**Methodology / Principal Findings:** We invited *T. cruzi* Ab reactive blood donors to enroll in the study from October 2018 to November 2019 with a follow up visit two years later. Blood samples were tested for *T cruzi* Ab by: Chagatest ELISA Lisado and Chagatest ELISA Recombinante v.4.0 (Wiener Lab, Argentina); VITROS Immunodiagnostic Products Anti-T.cruzi (Chagas) (Ortho-Clinical Diagnostics Inc., UK), and Architect Chagas (Abbott Laboratories, Germany). Target capture PCR was performed on lysed whole blood samples from enrollment visits and electrocardiograms on second visits.

Four hundred fifty donors were recruited, but 68 were excluded due to negative results on all study Ab assays. Ab level distributions were bimodal and classified as “high” or “low” at a calculated threshold for each of four assays. There were 160 donors with low and 179 with high Ab results on all assays. The remainder 43 were discordant reactive. Ninety-seven percentage of the PCR positive donors were among the concordant high Ab group. During the 2-4 year follow-up interval, relative Ab declines by three assays were significantly greater among those classified as low Ab and with negative PCR results.

**Conclusions / Significance:** Ab reactivity is associated with PCR-detectable parasitemia. Greater Ab declines were detected among donors with low and/or discordant Ab reactivity and negative PCR results, suggesting spontaneous parasite clearance in these donors.

## Introduction

Chagas disease (CD), caused by the *Trypanosoma cruzi* (*T. cruzi)* parasite, is a significant public health concern affecting approximately 6 million people worldwide [1]. Recognized as a neglected tropical disease, CD is primarily transmitted through contact with the feces of infected triatomine bugs [1]. Approximately 70 million people reside in areas at risk of exposure to this vector and parasite [2].

The diagnosis of chronically infected individuals relies on antibody (Ab) detection, as parasitemia is typically low and intermittent [1]. Due to well documented risks of transfusion transmission of *T. cruzi,* all blood donations are screened for *T. cruzi* Ab in endemic countries in Latin America, while donations from first time donors or travelers to endemic countries are screened for *T. cruzi* Ab in many non-endemic countries [3]. However, nearly one-third of individuals including blood donors identified as seropositive exhibit low Ab levels or discordant test results [4]. These borderline or low level Ab cases share risk factors with unequivocal seropositive cases, strongly suggesting true exposure to *T. cruzi* rather than false-positive results [5,6]. Since *T. cruzi* infection is widely considered to be lifelong [7], these low reactive samples are often considered as indicative of active infection and these patients and donors are deferred and counseled as infected and may be treated. This underscores the need to enhance the sensitivity of existing tests and forms the basis for PAHO’s recommendation of parallel screening using two different immunoassays for CD diagnosis [8].

Our previous study conducted in the highly endemic Chaco province of Argentina revealed a bimodal distribution of Ab signals in seroreactive blood donors when tested in parallel with six different immunoassays [6,9]. The bimodal distribution led us to hypothesize the existence of two infection outcome phenotypes, with low-level Ab cases reflecting spontaneously resolved and high-level Ab reflecting chronic/active *T. cruzi* infections. We further hypothesized that parasite clearance would diminish the antigenic stimulus, resulting in gradual seroreversion on follow-up testing. Consistent with our hypotheses, spontaneous cure has also been observed in other studies [10–15]

In the current study, we enrolled Chagas Ab-reactive donors at the same blood bank in the Chaco region as our previous study and conducted baseline and follow-up visits to confirm the bimodal distribution and correlate Ab levels with PCR results. Additionally, the follow-up visits allowed us to assess Ab decline over time and disease penetrance as measured by electrocardiogram (ECG).

## Methods

### Study design

In Argentina, blood donor screening policies mandate performance of two *T. cruzi* Ab tests based on different assay designs, such as parasite lysate and recombinant antigens. This prospective cohort study involved seropositive blood donors with at least one reactive screening test for *T. cruzi* at the Servicio Especializado en Hemoterapia in Chaco Province, Argentina, from 2009 to 2018. Eligible Ab-reactive donors were identified using blood bank records, and basic demographic data were retrieved. We organized the work within the province to secure local collaboration, ensuring an efficient process of recruitment, consent, sample acquisition and processing, and ECG testing. A local telemarketer was trained to make calls to donors in accordance with a protocol established with the local health providers.

The initial enrollment appointments and visits were conducted from October 2018 to November 2019. Donors completed a questionnaire regarding their risk factors for exposure to CD, and blood samples were collected for serological tests and PCR. Donors who reported previous treatment for CD were excluded from the study. The second follow-up visits took place from November 2021 to March 2023. During these visits, a second blood sample was collected for Ab testing, and each participant underwent an ECG.

### Serology testing

Four commercial tests were used for the characterization of *T. cruzi* Abs in the samples: Chagatest ELISA Lisado (Wiener Lab, Rosario, Argentina), Chagatest ELISA Recombinante v.4.0 (Wiener Lab, Rosario, Argentina), VITROS Immunodiagnostic Products Anti-T.cruzi (Chagas) (Ortho-Clinical Diagnostics Inc., Pencoed,Bridgend, UK), and Architect Chagas (Abbott Laboratories, Wiesbaden, Germany). The Chaco Province blood bank performed the Wiener and Abbott assays, and Vitalant Research Institute (San Francisco CA) was responsible for the Ortho assay. All *T. cruzi* Ab testing was performed according to the manufacturer’s instructions.

### Polymerase Chain Reaction (PCR)

During the enrolment appointment, 20 mL of EDTA-anticoagulated whole blood was drawn from each donor and mixed with an equal volume of guanidine (6 M)/ EDTA (200 mM) solution. Aliquots were prepared and stored at −20 degrees C until they were shipped to Vitalant Research Institute (San Francisco, CA) for PCR testing. Aliquots of lysed whole blood samples were tested using a target-capture (TC) real-time (RT) PCR assay, as previously described [12]. Capture of *T. cruzi* DNA was performed using magnetic beads coated with three 20-mer capture oligonucleotides: TCZ 1 CGAGCTCTTGCCCACACGGGAAAAAAAAAAAAAAAAAAAAAAAAAA; TCZ 2 CCTCCAAGCAGCGGATAGTTCAGGAAAAAAAAAAAAAAAAAAAAAAAAAA and; TCZ 3 TGCTGCASTCGGCTGATCGTTTTC-GAAAAAAAAAAAAAAAAAAAAA AAAAAA.

The captured DNA targets were eluted from the magnetic beads and real-time PCR amplified on an Applied Biosystems 7500 thermocycler. Briefly, 25 µL of DNA was added to 50 µL of PCR reaction mix. The PCR conditions were 10 min at 95° C, followed by 45 cycles of 30 sec at 95° C, 30 sec at 64° C, and 45 sec at 72° C. After completion of thermal cycling and real-time monitoring of cyber green intercalation, a dissociation step was performed, and the melting curves were analyzed. Product dissociations with one or two peaks at 80-82 degrees C were considered positive if the cycle threshold (CT) was less than 45 cycles. Eight replicate assays were performed, and the final interpretation was considered positive if at least two replicates produced a specific PCR product based CT and dissociation analysis.

### Electrocardiogram (ECG)

Standard 12-lead ECG was obtained using an electrocardiograph manufactured by Tecnologia Eletrônica Brasileira (São Paulo, Brazil)—model TEB ECGPC. All ECGs were transmitted to an ECG reading center at the Telehealth Center of the University Hospital of the Federal University of Minas Gerais, for standardized measurement, reporting and codification according to the Minnesota coding criteria (MC) in validated ECG data management software [16]. A certified cardiologist reviewed the exam, and a clinical report was sent back for counseling. All exams were manually codified according to the Minnesota code as normal or with minor or major electrocardiographic alterations, as previously described and validated for Chagas disease [17].

### Statistical analysis

Ab results were reported as signal-to-cutoff (S/CO) ratios, a function of the quantity/avidity of *T. cruzi* Ab present in samples. The results from our previous study in the same Argentinian Chaco region showed bimodal distributions of anti-*T. cruzi* antibodies in blood donors [6]. This bimodality suggests different host-parasite trajectories in the two groups, potentially reflecting high *versus* low or absent parasite burden and consequent CD pathogenicity. As such, we aimed to categorize all four serologic assays into “low” and “high” Ab levels in order to classify donor participants into two discrete groups based on their Ab reactivity. We fit mixture models, assuming underlying bimodal normal distributions. We then selected thresholds for each assay to optimally separate the two distributions using an “expectation minimization” algorithm [18] available at http://marcchoisy.free.fr//fmm/index.html.

Next, we explored differences in infection characteristics between high- and low-Ab reactive donor participants. In order to test the hypothesis that individuals with high Ab levels have higher rates of parasite persistence and parasite loads than those with low Abs, we compared the proportion with positive *T. cruzi* TC-PCR results at visit one across these groups.

We also compared the change in Ab reactivity between visit 1 and visit 2 according to baseline Ab category (high versus low) and TC-PCR results. The change in S/CO values for each serology test was defined as the difference between S/CO at follow-up and first enrolment visits, with negative values indicating a falling S/CO value. We analyze both relative and absolute change in S/CO. Relative S/CO values were defined as: (Follow-up S/CO value – Enrollment S/CO)/Enrollment S/CO value). A non-parametric significance test for continuous variables quantified Ab declines.

The presence of ECG abnormalities was evaluated as their distributions in three groups: high Ab levels, low Ab levels, and negative Ab subjects, considering as negative those individuals non-reactive for all serology tests at enrollment.

Analyses were conducted in R statistical software.

### Ethics statement

The study protocol was approved by the local Ethics committee at *Hospital Julio C. Perrando*, located in Resistencia City, the capital of Chaco province. All donor participants provided written informed consent before enrollment and follow-up visits.

## Results

### Cohort characteristics

A total of 455 donors participated in the first visit, with 450 providing valid results for all serological tests and valid PCR results. In the second visit, 390 of these 450 donors (86%) returned. Of those, 314 had their ECGs performed through the Telehealth system, allowing their ECG data to be entered and analyzed by the central reading core. The remaining 76 donors had their ECGs performed locally and the data could not be used in the study.

Table 1 summarizes the epidemiological characteristics of the 450 informative donors. The majority had significant epidemiological risk exposure: 358 donors (79.5%) had lived in a house where the vector of T. cruzi was present, 370 (82.2%) had lived in a house with mud walls, and 227 (50.4%) knew that a relative had suffered from Chagas Disease.

**Table 1.**
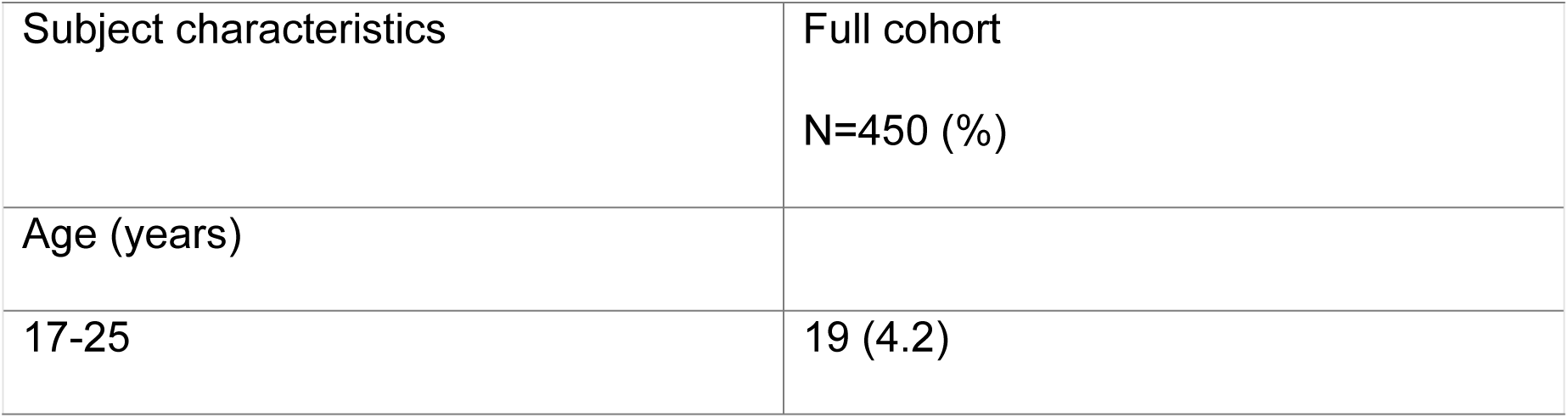
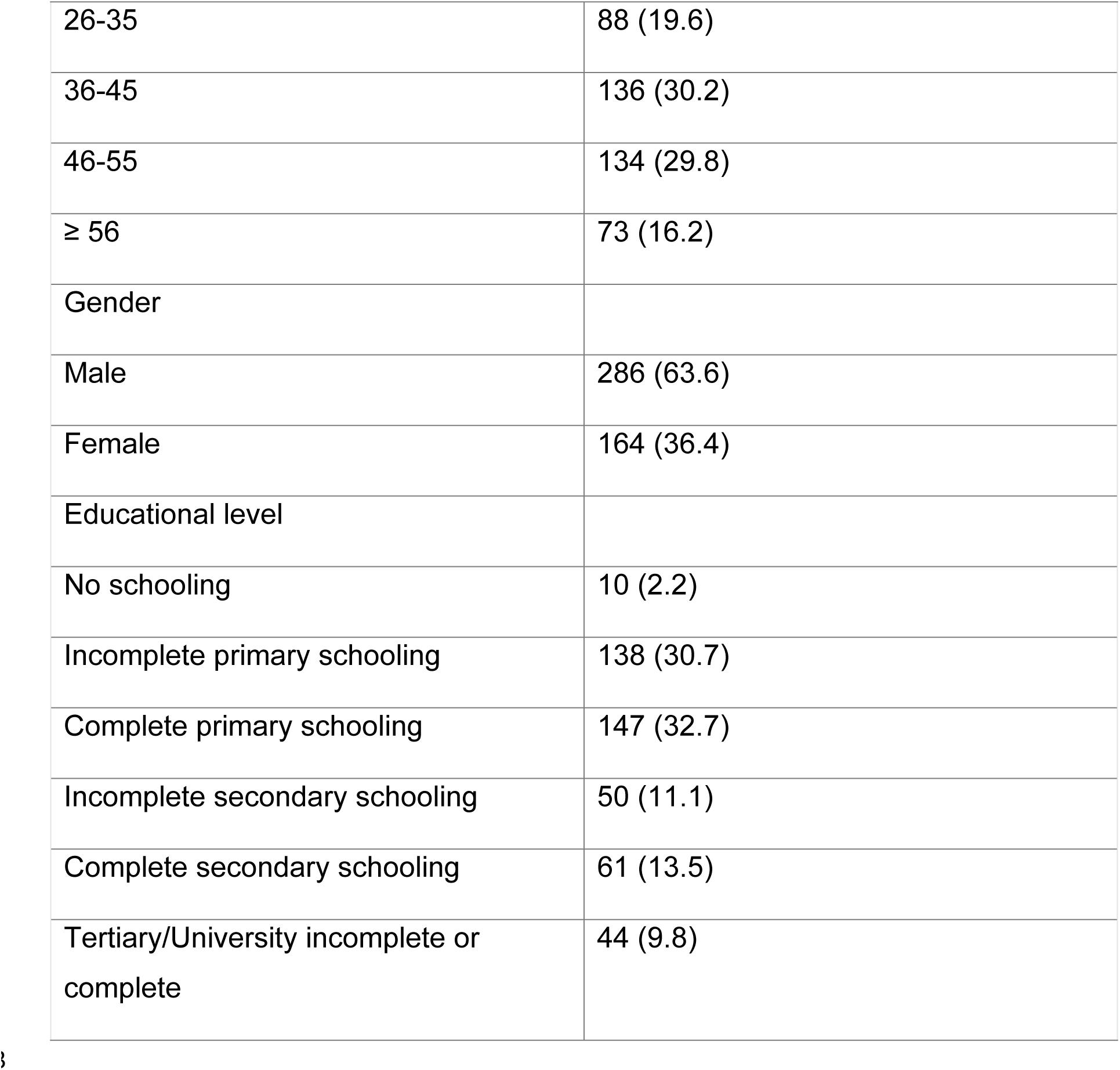
Characteristics of 450 enrolled *T. cruzi* seroreactive blood donors, Chaco region, 2018-2019.

### Serology testing and PCR detection

Serology analysis of the first visit samples revealed that 289 donors (64%) tested reactive on all four tests, 36 (8.0%) on three tests, 29 (6.4%) on two tests, and 28 (6.2%) on one test. The remaining 68 samples (15%) were negative on all four tests, and these were presumed to be false positive results in the initial donation screening; these 68 donors were excluded from subsequent analyses.

Fig 1 presents the distributions of S/CO values for each assay, revealing a clear bimodal distribution for each assay. Cut-off S/C values could be established to classify the samples into high and low Ab levels: 9.6 for the Architect Abbott assay, 5.2 for the Vitros assay; 4.8 for Wiener Lysate EIA and 4.7 for Wiener Recombinant EIA. There were 160 concordant (all four assays) low-level reactive samples and 179 concordant high-level Ab reactive samples. The remaining 43 samples were discordant with respect to low- and high-level reactivity on the four assays.

**Fig 1.**
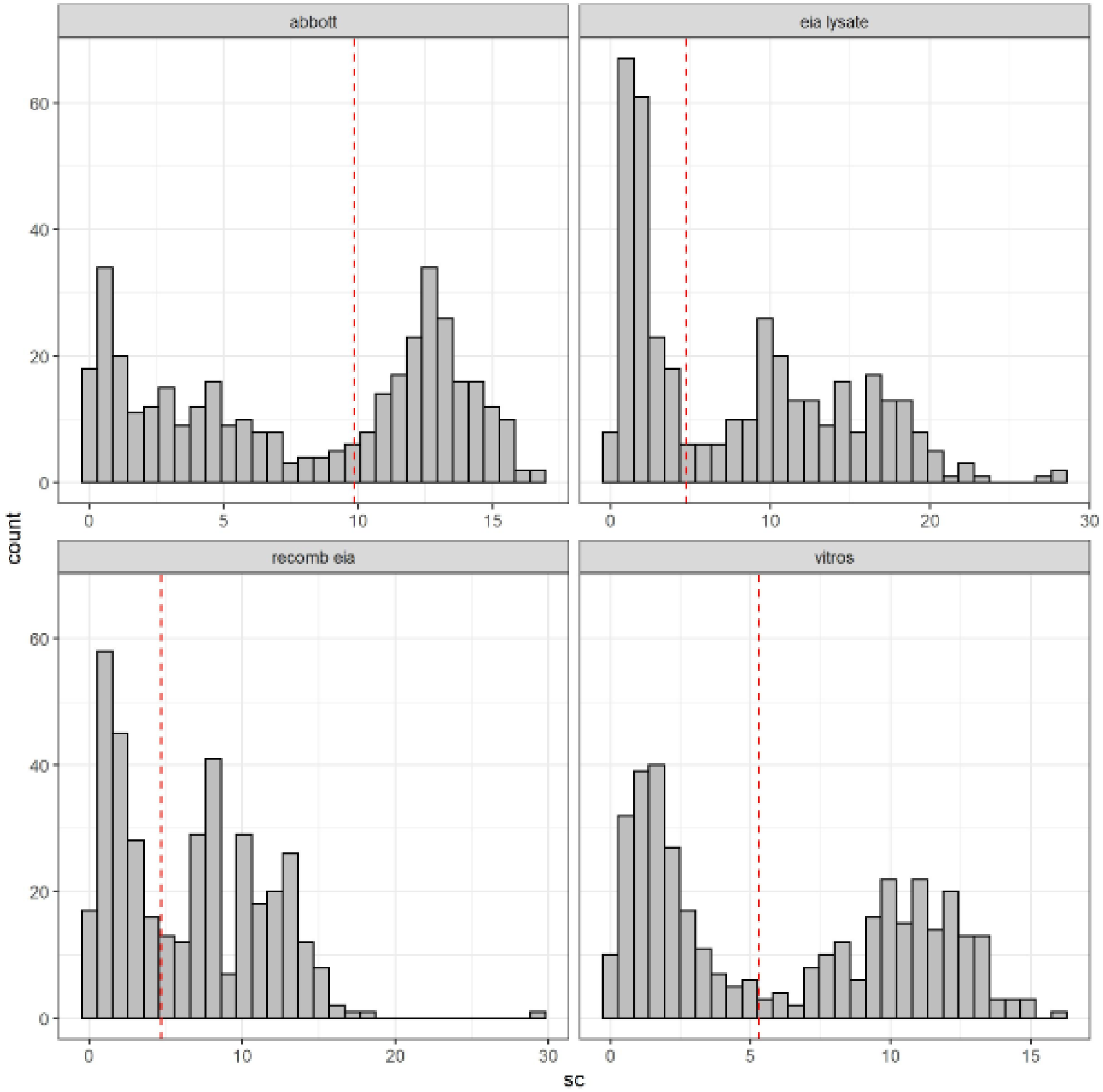
Determination of thresholds of bimodal signal-to-cutoff (S/CO) values distribution for each antibody test. Abbott: CMIA Chagas Architect, Abbott, Germany; EIA Lysate: ELISA Lisado, Wiener Lab., Argentina; Recomb EIA: ELISA Recombinante, Wiener Lab., Argentina; Vitros: Vitros Immunodiagnostics Products Anti-*T. cruzi* (Chagas) Assay (Ortho Clinical Diagnosis, Raritan NJ, USA. The dotted lines indicate the threshold values obtained using expectation minimization, assuming normal latent distributions. They resulted as 9.6 S/CO for CMIA, Architect Abbott; 5.2 S/CO for Vitros assay; 4.8 S/CO for Lysate EIA and 4.7 S/CO for Recombinant EIA.

Fig 2 demonstrates that this classification based on Ab reactivity levels correlates with the ability to detect parasitemia by TC-PCR. TC-PCR-positive results were associated with samples with high Ab levels, regardless of the assay used.

**Fig 2.**
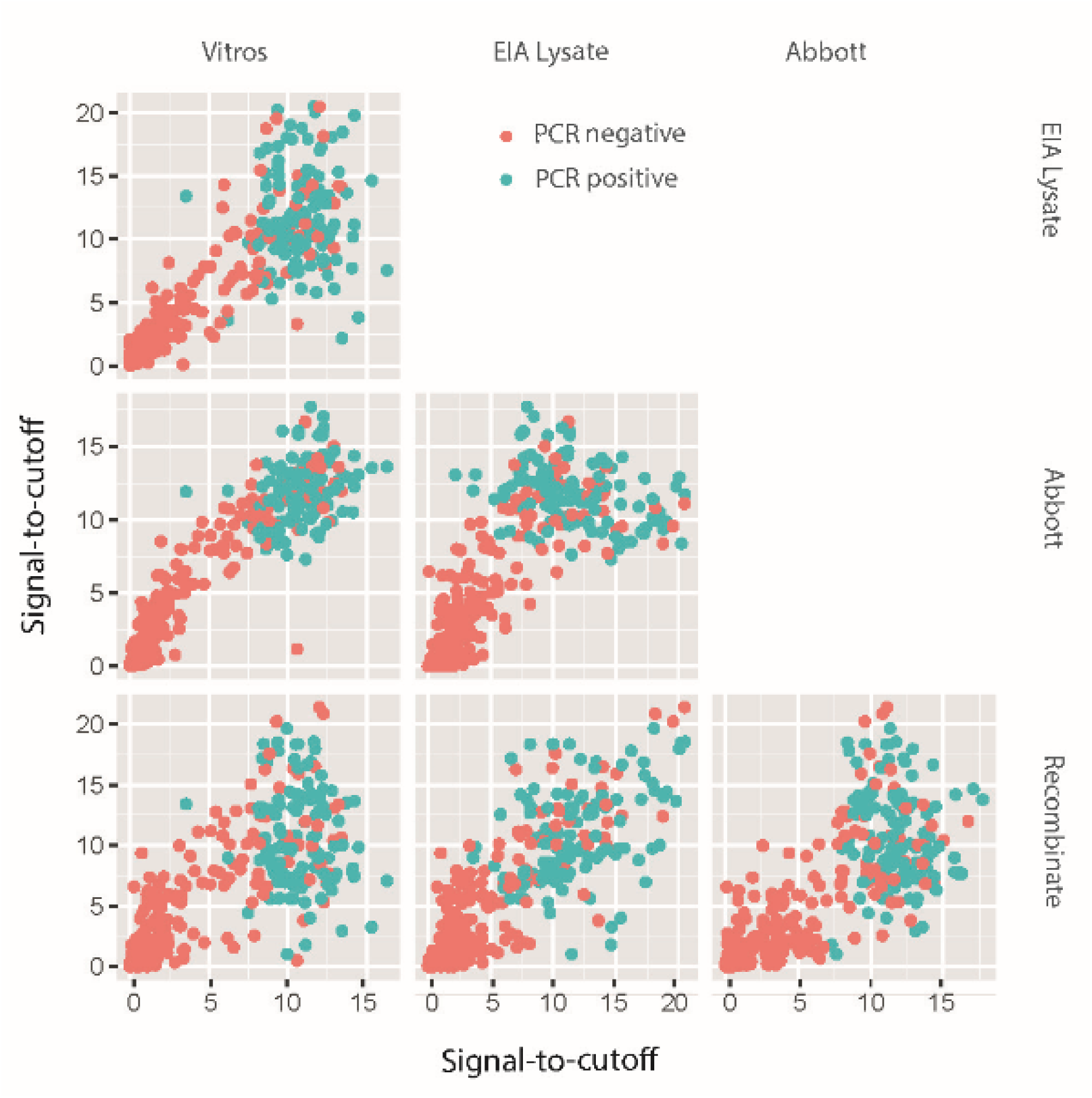
Correlation of antibody levels at visit 1 with T. cruzi PCR results. Scatter plots of signal-to-cutoff values at visit1. Abbott: CMIA Chagas Architect, Abbott, Germany; EIA Lysate: ELISA Lisado, Wiener Lab., Argentina; Recomb EIA: ELISA Recombinante, Wiener Lab., Argentina; Vitros: Vitros Immunodiagnostics Products Anti-*T. cruzi* (Chagas) Assay (Ortho Clinical Diagnosis, Raritan NJ, USA.

TC-PCR results were positive on 58-67% of the samples classified as high-level Ab reactive but only 0-1% of those classified as low-level Ab reactive, depending on the assays (Table 2). There were only four samples classified as low-level reactive by at least one of the assays that were PCR positive.

**Table 2.**
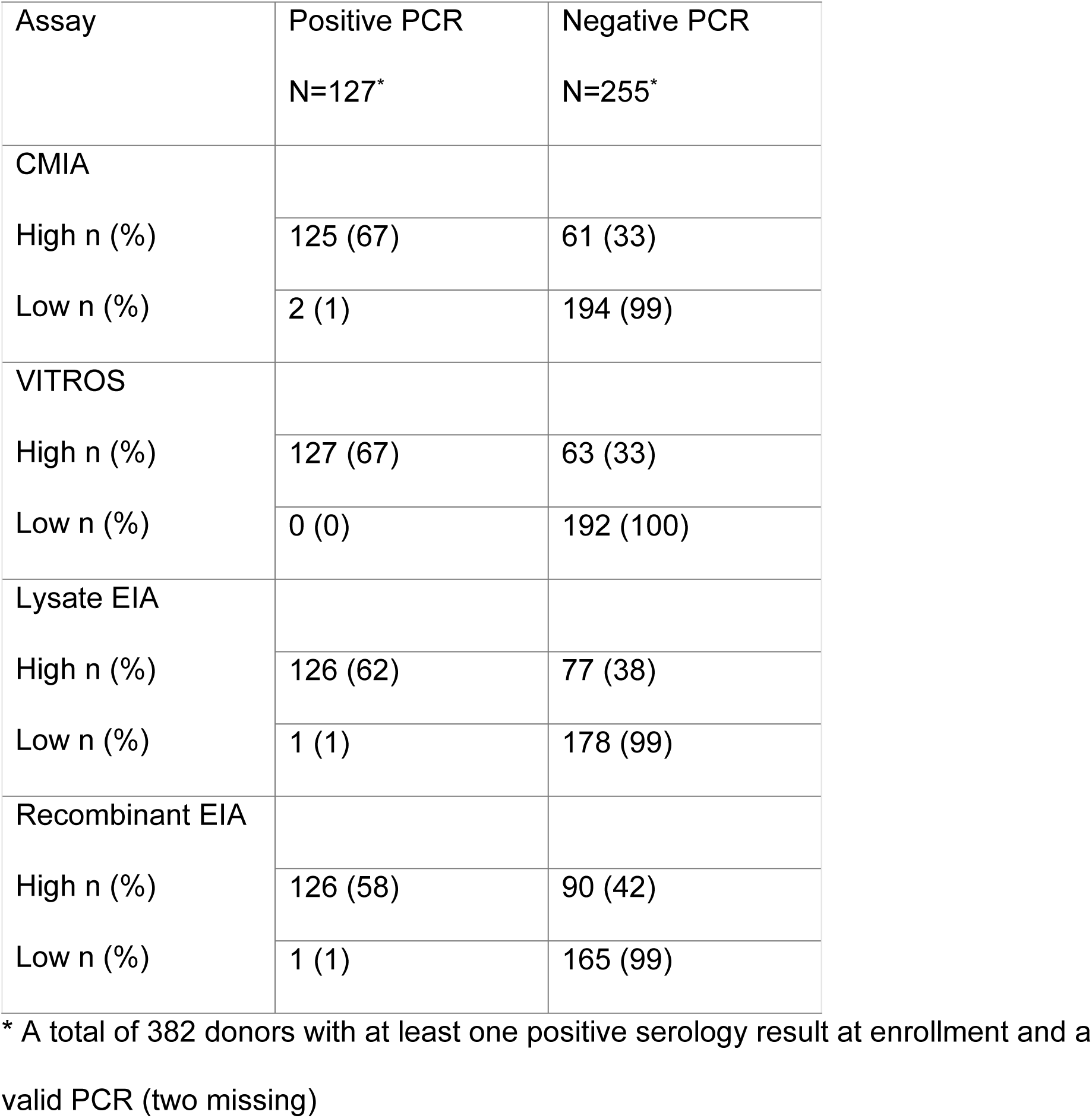
*T. cruzi* PCR samples distribution according to classification of high and low antibody levels for each serology test.

CMIA: Chagas Architect, Abbott; VITROS: VITROS Immunodiagnostics Products Anti-*T.cruzi* (Chagas) Assay (Ortho Clinical Diagnostics, Raritan NJ; Lysate EIA: Elisa lisado, Wiener Lab; Recombinant EIA: Elisa Recombinante, Wiener Lab.

### Ab levels on follow up relative to enrollment samples

The follow-up samples were collected 644 to 1511 days (median 899 days) after the enrollment samples. The relative decline in Ab reactivity was significantly higher among low-reactive samples when measured by three of the four kits used: the recombinant EIA and the two chemiluminescence assays (Fig 3). Measuring absolute decline was unable to detect differences (Fig 1 supplement).

**Fig 3.**
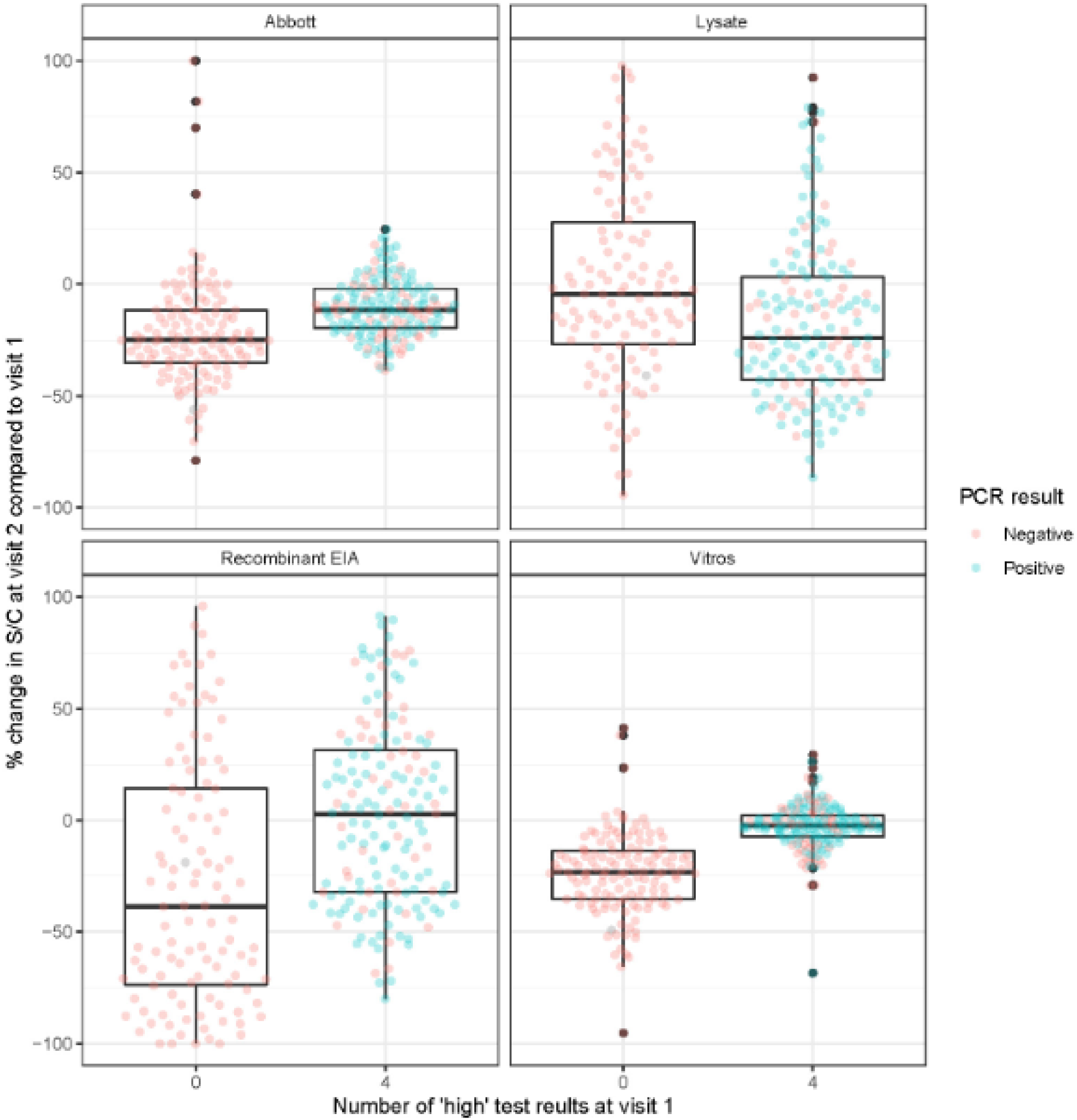
The relative decline in antibody levels over time. Box plot graphics show the relative percentage change in S/CO values at follow-up (visit 2) visit compared to enrollment (visit 1). The change in S/CO values for each serology test was defined as the difference between S/CO at follow-up and first enrolment visit, such as negative values indicating a falling S/CO value. For comparisons, 160 and 179 samples were fully concordant with four low (named as 0) and four high antibodies (named as 4) results, respectively. The blue dots represent PCR-positive samples, and the pink dots represent PCR-negative samples.

### Ab levels and ECG abnormalities

The presence of ECG abnormalities commonly associated with Chagas cardiomyopathy, such as prolonged QRS complex duration (>120 ms) and Right Bundle Branch Block (RBBB), was low in these asymptomatic donors. We could not establish an association between these abnormalities and Ab levels or PCR results (Table 3).

**Table 3.** Electrocardiogram abnormalities distribution in groups of donors with four tests concordant serology reactivity, as high and low antibody levels.

| **Serology results at enrollment** | **Four high serology tests** | **Four low serology tests** | **All negative serology tests** |
| --- | --- | --- | --- |
|  | N(%) | N(%) | N(%) |
| **QRS duration** |  |  |  |
| > 120 ms | 14 (11) | 10 (9) | 0 (0) |
| < 120 ms | 116 (89) | 102 (91) | 39 (100) |
| **RBB** |  |  |  |
| Yes | 6 (5) | 5 (4) | 0 (0) |
| No | 124 (95) | 107 (96) | 39 (100) |
| **Typical ECG abnormality** |  |  |  |
| Yes at least one | 29 (22) | 21 (19) | 4 (10) |
| No | 101 (78) | 91 (81) | 35 (90) |

RBBB: Right Bundle Branch Block

## Discussion

In this study, we established a prospective cohort of *T. cruzi* seroreactive blood donors from a highly endemic area in the Chaco region of Argentina. We confirmed a bimodal distribution of Ab levels and demonstrate for the first time that Ab reactivity levels correlate with parasite detection in lysed whole blood by a sensitive TC-PCR assay.

These findings align with those from our previous cross-sectional study of blood donors from the same region, in which we described a bimodal distribution of Ab S/CO values on several *T. cruzi* Ab screening tests; however, *T. cruzi* parasitemia and longitudinal Ab reactivity patterns were not evaluated in that study [6]. In the current study, we have demonstrated that the biological process likely underlying the bimodal distribution is persistence of *T. cruzi* infection based on parasitemia detected by TC-PCR. Furthermore, we were able to establish a threshold S/CO value for each of the four assays for classifying cases into high and low Ab level groups that highly correlated with parasite persistence by PCR and can be used for counselling and decisions on treatment.

The association between PCR and Ab levels was initially described in a study we conducted with blood donor samples from Honduras, the USA, and Brazil [12]. However, due to the pre-screening process of the samples that excluded low reactive samples in that study, a bimodal distribution of Abs was not demonstrated.

Despite the relatively short interval between enrolment and follow-up visits in the current study (median 899 days), Ab decline was observed in the low reactive group but not in the high reactive group by three of the four assays. This suggests that individuals in the low Ab group had cleared or were very effectively controlling parasitemia and replication in tissue reservoirs, reducing the antigenic stimulus, which lead to Ab waning and eventually complete seroreversion.

Notably, 93 samples showed discrepant results (reactive to only 1, 2, or 3 assays), representing approximately 50% of the low reactive samples. Discrepant test results are common in donor screening and Chagas disease diagnosis, and are often viewed as due to a lack of sensitivity in the assays. Conversely, our data suggest that these discrepancies indicate spontaneous cure or effective control of parasite replication, implying that parallel screening with two immunoassays to detect discrepant cases may not be necessary.

According to PAHO, less than 10% of T. cruzi infected individuals receive timely diagnosis and treatment [19]. Simplifying and optimally eliminating the requirement for a parallel testing algorithm would improve access to diagnosis in low-income areas.

Another important point is that most clinical trials for Chagas disease rely on PCR results for the initial inclusion criteria of subjects [20]. However, *T. cruz*i PCR is generally challenging to interpret due to low and intermittent parasitemia, requiring multiple replicates that often lead to discrepant results. By establishing a cut-off value using different serological tests, we can improve the screening process for clinical trials of therapeutics, as the proportion of PCR-positive cases is much higher among those with elevated Ab levels.

Ab levels have recently been associated with the development of Chagas cardiomyopathy [21, 22]. In the present study, we could not establish an association between disease penetrance and Ab levels, likely due to the young age of our cohort and short follow-up period following asymptomatic blood donations.

In summary, this study provides additional evidence for the bimodal distribution of Ab reactivity in *T. cruzi-*exposed donors/patients, and the correlation with a sensitive PCR assay results suggests that spontaneous cure may be responsible for low reactivity and discordant Ab results. We have also established potential serological cut-off values that could help classify donors/patients who are more likely to have persistent parasitemia, which could be used for prognosis and to indicate the need for treatment.

## Data Availability

The data that support the findings of this study are publycly available from Figshare with the identifier 10.6084/m9.figshare.27897405

## Acknowledgments

We acknowledged technical and laboratory personnel at *Centro Especializado en Hemoterapia del Chaco*. We appreciate the administrative personnel for dedicated work in recruiting blood donors at blood center. We thank the M o H of the Chaco Province for the relevant support.

## Supporting information

S1 Fig. Absolute changes in antibody levels

**Figure 1 supplemental:**
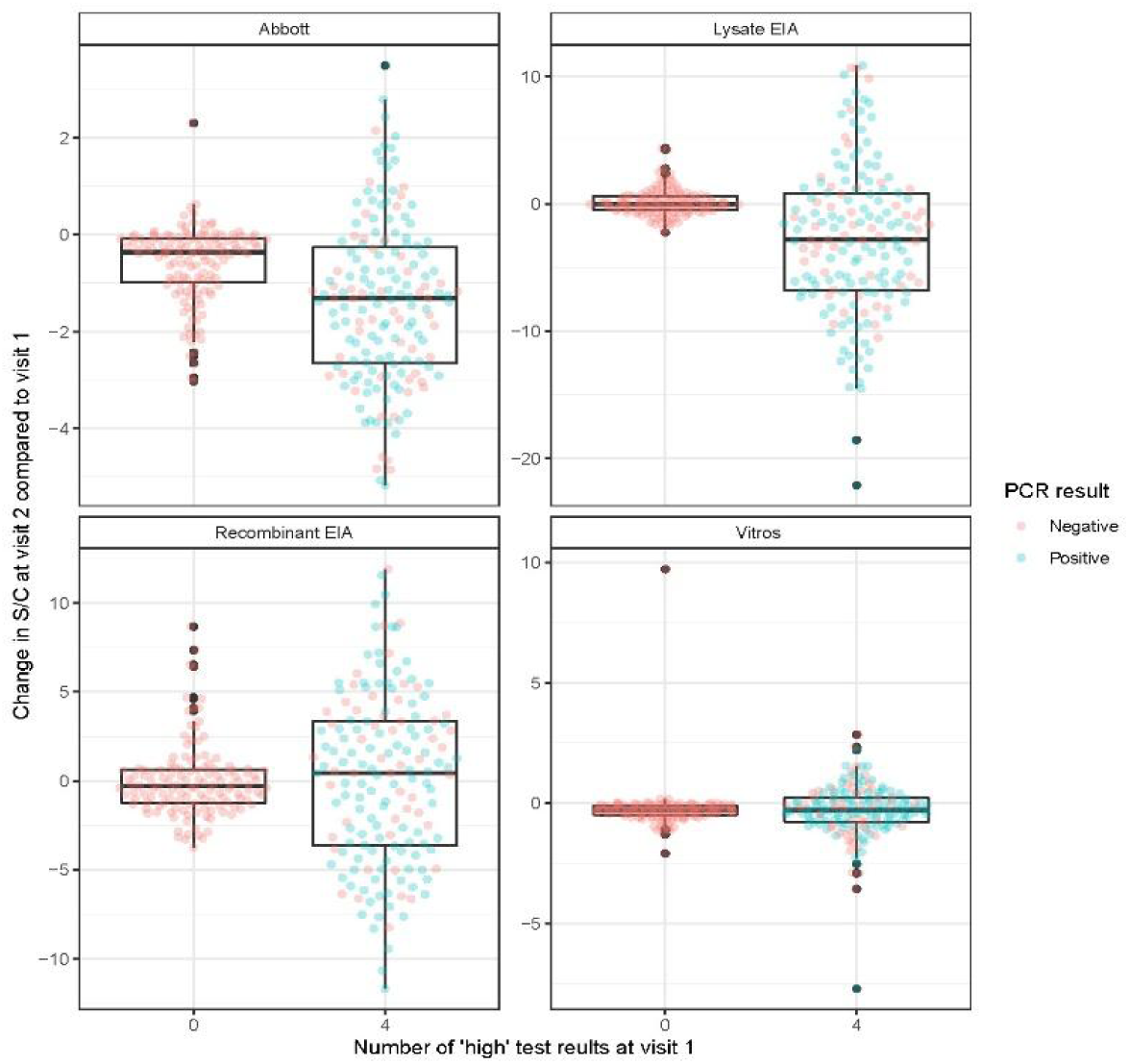
Absolute changes in antibody levels.

## References

1. Sabino EC, Nunes MCP, Blum J, Molina I, Ribeiro ALP. Cardiac involvement in Chagas disease and African trypanosomiasis. Nat Rev Cardiol [Internet]. 2024 Jul 15; Available from: 10.1038/s41569-024-01057-3

2. Pérez-Molina JA, Molina I. Chagas disease. Lancet. 2018 Jan 6;391(10115):82–94. doi: 10.1016/S0140-6736(17)31612-4.

3. Crowder LA, Wendel S, Bloch EM, O’Brien SF, Delage G, Sauleda S, et al. WP-TTID Subgroup on Parasites. International survey of strategies to mitigate transfusion-transmitted Trypanosoma cruzi in non-endemic countries, 2016-2018. Vox Sang. 2022 Jan;117(1):58-63. doi: 10.1111/vox.13164.

4. Sabino EC, Salles NA, Sarr M, Barreto AM, Oikawa M, Oliveira CD, et al. Enhanced classification of Chagas serologic results and epidemiologic characteristics of seropositive donors at three large blood centers in Brazil. Transfusion. 2010 Dec;50(12):2628–37. doi: 10.1111/j.1537-2995.2010.02756.x.

5. Salles NA, Sabino EC, Cliquet MG, Eluf-Neto J, Mayer A, Almeida-Neto C, et al. Risk of exposure to Chagas’ disease among seroreactive Brazilian blood donors. Transfusion. 1996 Nov;36(11-12):969–73. doi: 10.1046/j.1537-2995.1996.36111297091740.x.

6. Remesar M, Sabino EC, Del Pozo A, Mayer A, Busch MP, Custer B. Bimodal distribution of Trypanosoma cruzi antibody levels in blood donors from a highly endemic area of Argentina: what is the significance of low-reactive samples? Transfusion. 2015 May 27;55(10):2499–504. doi: 10.1111/trf.13180.

7. de Sousa AS, Vermeij D, Ramos AN Jr, Luquetti AO. Chagas disease. Lancet [Internet]. 2023 Dec 7; Available from: 10.1016/S0140-6736(23)01787-7

8. Pan American Health Organization. Guidelines for the Diagnosis and Treatment of Chagas Disease. Washington, DC: PAHO; 2019.

9. Remesar MC, Gamba C, Colaianni IF, Puppo M, Sartor PA, Murphy EL, et al. Estimation of sensitivity and specificity of several Trypanosoma cruzi antibody assays in blood donors in Argentina. Transfusion. 2009 Nov;49(11):2352–8. doi: 10.1111/j.1537-2995.2009.02301.x.

10. Francolino SS, Antunes AF, Talice R, Rosa R, Selanikio J, de Rezende JM, et al. New evidence of spontaneous cure in human Chagas’ disease. Rev Soc Bras Med Trop. 2003 Apr 22;36(1):103–7. doi: 10.1590/s0037-86822003000100014.

11. Dias JCP, Dias E, Martins-Filho OA, Vitelli-Avelar D, Correia D, Lages E, et al. Further evidence of spontaneous cure in human Chagas disease. Rev Soc Bras Med Trop. 2008 Sep-Oct;41(5):505–6. doi: 10.1590/s0037-86822008000500014.

12. Sabino EC, Lee TH, Montalvo L, Nguyen ML, Leiby DA, Carrick DM, et al. Antibody levels correlate with detection of Trypanosoma cruzi DNA by sensitive polymerase chain reaction assays in seropositive blood donors and possible resolution of infection over time. Transfusion. 2012 Sep 25;53(6):1257–65. doi: 10.1111/j.1537-2995.2012.03902.x.

13. Bertocchi GL, Vigliano CA, Lococo BG, Petti MA, Viotti RJ. Clinical characteristics and outcome of 107 adult patients with chronic Chagas disease and parasitological cure criteria. Trans R Soc Trop Med Hyg. 2013 Jun;107(6):372–6. doi: 10.1093/trstmh/trt029.

14. Viotti R, Vigliano C, Lococo B, Bertocchi G, Petti M, Alvarez MG, et al. Long-term cardiac outcomes of treating chronic Chagas disease with benznidazole versus no treatment: a nonrandomized trial. Ann Intern Med. 2006 May 16;144(10):724–34. doi: 10.7326/0003-4819-144-10-200605160-00006.

15. Zeledón R, Dias JC, Brilla-Salazar A, de Rezende JM, Vargas LG, Urbina A. Does a spontaneous cure for Chagas’ disease exist? Rev Soc Bras Med Trop. 1988 Jan-Mar;21(1):15–20. doi: 10.1590/s0037-86821988000100003.

16. Pinto-Filho MM, Paixão GM, Gomes PR, Soares CPM, Singh K, Rossi VA, et al. Electrocardiographic findings and prognostic values in patients hospitalised with COVID-19 in the World Heart Federation Global Study. Heart. 2023 Apr 12;109(9):668–73. doi: 10.1136/heartjnl-2022-321754.

17. Ribeiro ALP, Marcolino MS, Prineas RJ, Lima-Costa MF. Electrocardiographic abnormalities in elderly Chagas disease patients: 10-year follow-up of the Bambui Cohort Study of Aging. J Am Heart Assoc. 2014 Feb 7;3(1):e000632. doi: 10.1161/JAHA.113.000632.

18. Trang NV, Choisy M, Nakagomi T, Chinh NTM, Doan YH, Yamashiro T, et al. Determination of cut-off cycle threshold values in routine RT-PCR assays to assist differential diagnosis of norovirus in children hospitalized for acute gastroenteritis. Epidemiol Infect. 2015 Nov;143(15):3292–9. doi: 10.1017/S095026881500059X.

19. Apr 13. Less than 10% of those infected with Chagas disease receive timely diagnosis and treatment [Internet]. [cited 2024 Jul 24]. Available from: https://www.paho.org/en/news/13-4-2022-less-10-those-infected-chagas-disease-receive-timely-diagnosis-and-treatment

20. Bosch-Nicolau P, Fernández ML, Sulleiro E, Villar JC, Perez-Molina JA, Correa-Oliveira R, et al. Efficacy of three benznidazole dosing strategies for adults living with chronic Chagas disease (MULTIBENZ): an international, randomised, double-blind, phase 2b trial. Lancet Infect Dis. 2024 Apr;24(4):386–94. doi: 10.1016/S1473-3099(23)00629-1.

21. Nunes MCP, Buss LF, Silva JLP, Martins LNA, Oliveira CDL, Cardoso CS, et al. Incidence and Predictors of Progression to Chagas Cardiomyopathy: Long-Term Follow-Up of Trypanosoma cruzi-Seropositive Individuals. Circulation. 2021 Sep 27;144(19):1553–66. doi: 10.1161/CIRCULATIONAHA.121.055112.

22. Buss LF, Campos de Oliveira-da Silva L, Moreira CHV, Manuli ER, Sales FC, Morales I, et al. Declining antibody levels to Trypanosoma cruzi correlate with polymerase chain reaction positivity and electrocardiographic changes in a retrospective cohort of untreated Brazilian blood donors. PLoS Negl Trop Dis. 2020 Oct 27;14(10):e0008787. doi: 10.1371/journal.pntd.0008787.

